# Multi-omics Analysis Reveals Biomarkers that Contribute to Biological Age Rejuvenation in Response to Therapeutic Plasma Exchange

**DOI:** 10.1101/2024.08.02.24310303

**Authors:** Matias Fuentealba, Dobri Kiprov, Kevin Schneider, Wei-Chieh Mu, Prasanna Ashok Kumaar, Herbert Kasler, Jordan B. Burton, Mark Watson, Heather Halaweh, Christina D. King, Zehra Stara Yüksel, Chelo Roska-Pamaong, Birgit Schilling, Eric Verdin, David Furman

## Abstract

The rapid increase in the proportion of older adults worldwide poses a huge challenge for healthcare systems. Currently, age-related chronic diseases account for over 90% of annual healthcare expenditures (more than $4.1 trillion) in the US alone. Thus, new therapies to improve healthspan and reduce the burden of chronic disease are needed. Herein, we conducted a phase 3 controlled clinical trial to evaluate the biological age (BA) effects of different therapeutic plasma exchange (TPE) modalities in ambulatory individuals. We longitudinally profiled the subjects to measure changes in the epigenome, proteome, metabolome, glycome, and shifts in immune cell composition (cytomics). We demonstrate that administering TPE supplemented with intravenous immunoglobulin (IVIG) (TPE-IVIG) on a biweekly regime (two sessions in the first week, followed by a three-week break) is a robust therapy for BA rejuvenation. This intervention induced coordinated cellular and molecular ‘omics’ responses, reversed age-related immune decline, and modulated key cellular senescence-associated proteins. Integrative analysis revealed baseline biomarkers associated with successful responses, indicating that TPE-IVIG treatment benefits those with a poorer baseline health status. In summary, this is the first multi-omics study to examine the effectiveness of various TPE modalities, which demonstrate biological age rejuvenation and the molecular features associated with this rejuvenation.

## Introduction

Every country in the world is experiencing sustained growth in the proportion of older adults in its population. Current estimates predict that by 2030, almost a sixth of the world’s population will be aged 60 years or older^1^. As age is the single most significant risk factor for chronic disease, these population dynamics pose a huge challenge for healthcare systems. However, a person’s true age is much more than the chronological number of years they have lived. Factors like stress, diet, sleep, genetics, and infections influence the rate at which our bodies age, and biologically older individuals are more likely to develop diseases and experience premature mortality^2–8^. In recent years, we have witnessed an increasing number of studies focused on estimating a person’s biological age (BA) at the molecular level based on changes to epigenomics^9^, proteomics^3,10^, metabolomics^11^, and other ‘omics’ measurements^12,13^. The most accurate and well-established methods utilize changes in DNA methylation (DNAm) patterns that occur with age, followed by machine learning techniques to construct biological age (BA) predictors. These BA ‘epigenetic clocks’ are often predictive of diseases and mortality^12^, and sensitive to various lifestyle interventions. For instance, moderate weight loss (<5%) decreases epigenetic age by 1.1 years (Horvath clock), and individuals with more than 5% in weight loss show a decrease of 7.2 BA months in a 18-months period^14^. Nutrition also has a significant impact on methylation clocks. For example, methylation-supportive diets combined with lifestyle changes can reduce biological age by 4.6 years in females^15^ and 1.96 years in males^16^. Healthy dietary habits maintained for up to 2 years can reduce age-adjusted biological age by 0.66 years (GrimAge)^17^. The effects of diet and nutritional supplementation seem to depend on the study population. For instance, in overweight African Americans, vitamin D3 supplementation can decrease epigenetic age by approximately 1.9 years^18^ whereas in vitamin D-deficient subjects, it decreases epigenetic age by only 1.3 years^19,20^. Interestingly, a 60-day relaxation practice scheme can also significantly impact epigenetic age with a rejuvenation of 4.67 years in healthy subjects^21^. Pharmacological interventions have highlighted the potential of small molecules to reduce BA. For instance, we observed a reduction in epigenetic age acceleration by 2.77 years (Horvath clock) and 3.43 years (Hannum clock) in patients with diabetes mellitus undergoing metformin treatment^22^. Other pharmacological agents, such as dasatinib and quercetin (NCT04946383), and rapamycin (NCT04608448), are currently undergoing clinical trials for reducing BA^23^.

Methods to manipulate blood composition have proven promising in decreasing BA clocks and improving health status. For instance, intramuscular injection of human umbilical cord plasma concentrate into elderly human individuals has shown that youth factors substantially improve clinical biomarkers and reduce biological age by 0.82 years (using GrimAge)^24^. Several studies have shown that therapeutic plasma exchange (TPE), first studied in animals in 1914^25^, can also significantly improve the outcomes of various medical indications. TPE, first used to treat Macroglobulinemia in 1963^26^ has recently been given Food and Drug Administration Emergency Use Authorization for the treatment of COVID-19^27^. Strikingly, up to 65% of patients with long COVID have shown improved peripheral neuropathy, fatigue, stamina, and brain fog after TPE treatment^28^. Interestingly, in cases of yellow phosphorus poisoning, TPE showed strong beneficial effects such as removal of the poison and improved liver function with associated changes in secreted circulating proteins and metabolites^29^.

In the context of aging, we recently conducted a study of 8 individuals, demonstrating a remodeling of the immune system in the blood of older individuals and a decrease in proteins associated with aging after over five repeated TPE treatments^30^. Expanding on our previous findings, this study applies a multi-omics systems biology approach to longitudinally profile 30 individuals who underwent three therapeutic plasma exchange (TPE) modalities (10 individuals each): monthly TPE, biweekly TPE, and biweekly TPE combined with intravenous immunoglobulin (IVIG). We utilized BA deceleration as the primary endpoint by measuring 35 independent epigenetic clocks and estimated the BA rejuvenation effects caused by each intervention. Integrative analysis encompassing lipidomics, proteomics, metabolomics, cytomics, and glycomics, identified biomarkers in each ‘omics’ that are correlated with the responses to TPE and baseline clinical and ‘omics’ features that are predictive of the rejuvenation response to TPE treatment.

## Results

### TPE induces biological age rejuvenation and IVIG supplementation enhances the effect

Subjects underwent therapeutic plasma exchange in two temporal regimes. Ten individuals were subjected to two sessions during the first week, followed by three weeks with no sessions for 3 months (biweekly regime), and ten individuals received one session per month 6 times (monthly regime) (Figure 1A). Blood samples were taken in both regimes before sessions 1 (baseline), 4 and 6. In addition to TPE, we evaluated in ten individuals the effects of biweekly intravenous injections of immunoglobulin (IVIG) supplementation, which has previously been shown to enhance the immune system’s ability to fight infections^31,32^. The control group received sham plasma exchange, which mimicked TPE but without fluid replacement (see Methods).

**Figure 1.**
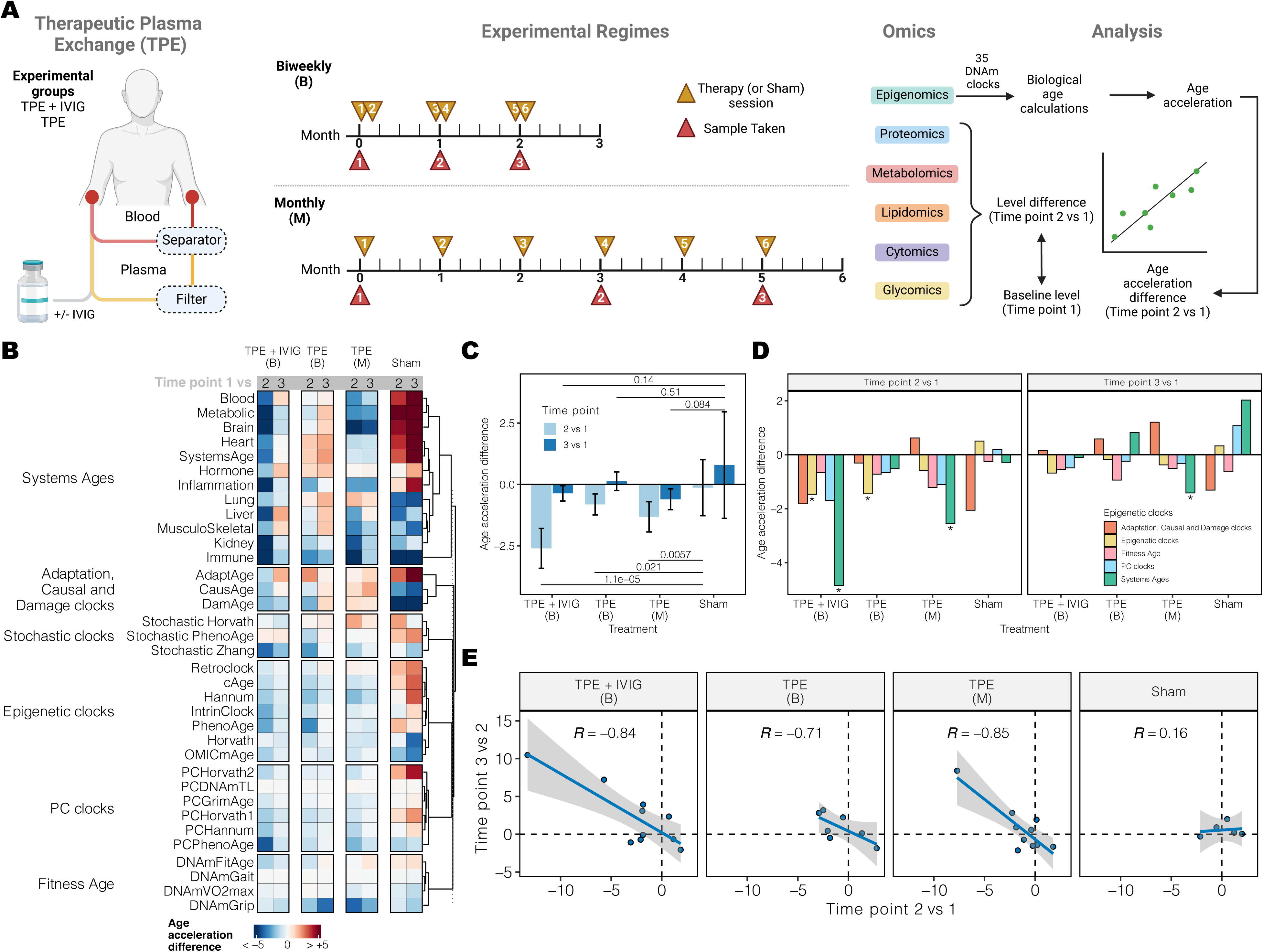
Therapeutic plasma exchange induces a coordinated biological age deceleration. A) Schematic representation of TPE treatment. In TPE, blood cells and plasma are separated. Plasma is then filtered and replaced with IVIG or fluids, and together with blood cells are returned to the patient’s circulation. Two temporal regimes were tested, with three blood samples taken per session. Samples were used to perform multi-omics profiling. Epigenomics data was used to calculate the differences in BA induced by the treatments. Correlation analyses were performed, comparing baseline and changing levels of the omics features and BA differences. B) Biological age estimations using 35 epigenetic clocks. In all groups, we calculated the difference between the age-adjusted biological age at time points 2 and 3 compared to time point 1 (age acceleration difference). C) Average age acceleration difference across epigenetic clocks. Error bars indicate the 95% confidence intervals of the mean. P-values at the top and bottom indicate the significance of the difference between the treatments and sham using a Wilcoxon test. D) Average age acceleration difference for different groups of epigenetic clocks (labels are consistent with panel B). Asterisks indicate age acceleration differences significantly different from zero. E) Comparison of average age acceleration difference between time point 2 vs 1 and time point 3 vs 2. Correlations were calculated using Pearson’s method.

We collected epigenetic data from the treated subjects and calculated DNA methylation age using 35 different epigenetic clocks. We adjusted the biological ages for each clock by the chronological age, resulting in a metric of age acceleration. This metric represents the deviation of BA from individuals of the same age. To estimate the effects on BA induced by the interventions, we calculated the difference between the age acceleration at time points 2 or 3 compared to baseline (time point 1). This age acceleration difference was negative if the interventions reduced biological age and positive if the interventions increased biological age, independently of age (Figure 1B). We observed that at time point 2, all interventions induced a negative age acceleration difference compared to sham (Figure 1C). TPE + IVIG treatment displayed the largest reduction in BA, with an average decrease of 2.61 years (p = 1.1e-05). In the case of TPE, the monthly regime showed a greater decrease in biological age with an average BA rejuvenation of 1.32 years (p = 5.7e-03), suggesting that more frequent sessions do not necessarily lead to a greater BA rejuvenation effect. Surprisingly, we observed no significant BA differences at time point 3 compared to sham in any group, suggesting potential compensatory mechanisms that mitigate the anti-aging effects after multiple sessions.

Given that epigenetic clocks capture different aspects of aging, we asked which type of BA clock had the greatest effect. To do so, we grouped these clocks into 6 different types, depending on the method and outcome regressed (see Methods). At time point 2, Systems age clocks^33^ showed the largest decrease in age-adjusted BA in the TPE + IVIG group (4.85 years, FDR = 2.52e-04) and the monthly TPE intervention (2.55 years, FDR = 9.90e-03) but not in the TPE biweekly group (FDR = 0.91) (Figure 1D). More traditional epigenetic clocks also showed significant age acceleration difference in the TPE + IVIG (FDR = 5.12e-03) and TPE (FDR = 1.75e-03) interventions under a biweekly regime. Consistent with the global calculations, none of the clock groups showed a significant difference in BA at time point 3 except for the monthly TPE intervention (FDR = 6.40e-03). Given the observation that at time point 3, the effects were largely attenuated, we wondered if this effect was dependent on the effects observed at time point 2, which could indicate a potential compensatory mechanism. Indeed, for all interventions, a strong negative correlation was observed between the age acceleration differences at time points 2 from baseline and those between time point 3 and 2 (Figure 1E). The sham group showed no such correlation. Overall, these results indicate that individuals with the most rejuvenation effects during the first three sessions tend to show a greater BA increase in subsequent sessions.

### Omics changes linked to rejuvenation in TPE + IVIG

To provide a better understanding of the molecular basis of the BA rejuvenation observed, we performed multi-omics profiling on the same samples, including cytomics, glycomics, lipidomics, metabolomics, and proteomics (see Methods). To identify omics markers associated with the rejuvenation effects observed in time point 2, we calculated the correlations between the age acceleration differences induced by each intervention and the change in levels of each feature for each omics. We focused on features whose correlation was significantly different from zero and the sham intervention (see Methods, Figure 2A).

**Figure 2.**
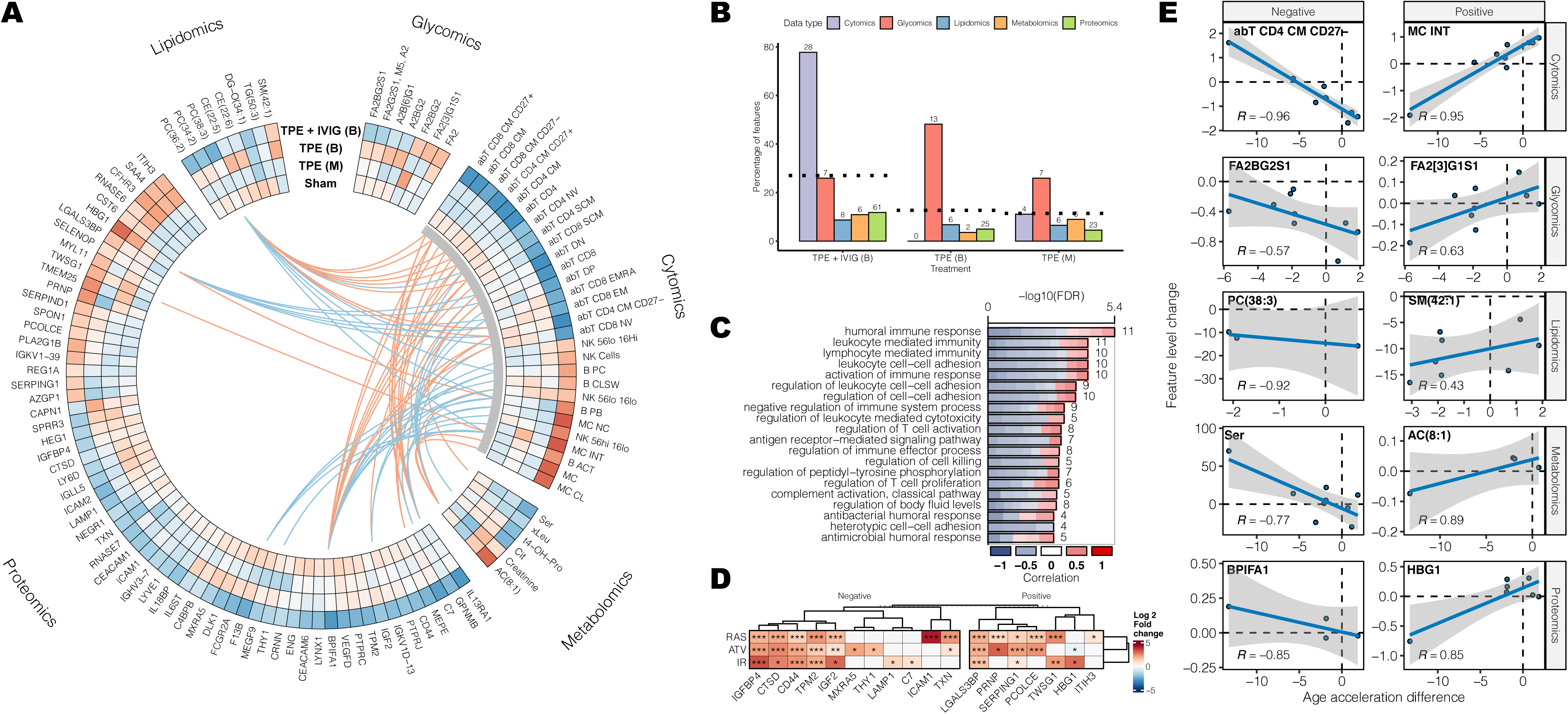
Multiple ‘omics’ features and cellular senescence proteins track with biological age rejuvenation response to TPE-IVIG. A) Correlations between the rejuvenation effects and changes in levels of omics features. We only display features significantly different from zero and sham in the TPE + IVIG group. Lines between the features at the center indicate inter-feature correlations between cell composition changes and other omics. B) Percentage of features significantly changing in each omics and group. The dotted line indicates the average across omics. C) Gene ontology enrichment analysis of proteins with levels correlated with the responses to TPE + IVIG. D) Change in abundance in senescent cells of proteins relevant for the TPE + IVIG response. E) Features with the strongest positive and negative correlation for each omics with the rejuvenation effects.

Overall, we observed that TPE + IVIG induced the most changes in omics profiles with nearly 77% of the cell types measured in the cytomics changing in proportion in coordination with the biological age differences (Figure 2B). However, we observed minor or no changes in cell types in TPE interventions, indicating that IVIG induced major changes in cell type composition, some of which could potentially contribute to the rejuvenation effects of BA. For example, we found that the BA rejuvenation effects were associated with a higher proportion of CD8 and CD4 naive T cells, a hallmark of immune aging, and lower levels of NK cells and monocytes. These results are consistent with previous observations that CD8 and CD4 naive T cells significantly drop with age, while NK cells and monocytes tend to increase with age^34,35^. Across interventions, glycomics showed the largest proportion of changes, with nearly a third (33.3%) of the glycans measured changing. In proteomics, 61 proteins, representing 11% of the proteome measured in this study, displayed significant correlations with the BA changes in TPE + IVIG, compared to less than 5% for the TPE group. To provide a better understanding of the function of these proteins, we performed gene ontology enrichment analysis. Proteins with levels correlated with rejuvenation effects in TPE + IVIG were mostly involved in activation of the immune response, complement cascade, and cell-cell adhesion (Figure 2C). We also analyzed whether the proteins that significantly correlated with the BA rejuvenation effects were also associated with cellular senescence. To do so, we compared this set of proteins changing with TPE against those in the SASP Atlas^36^. The SASP Atlas included proteins changing during senescence induced in fibroblasts by X-ray irradiation (genotoxic stress-induced), RAS overexpression (oncogene-induced) and Atazanivir treatment (treatment-induced). We observed a significant up-regulation of proteins correlated with rejuvenation effects in senescent fibroblasts, suggesting that the modulation of senescence may partially drive these rejuvenation effects (Figure 2D).

Given the large cell type composition changes observed, we wondered if these were correlated with other omics features that were significantly changing (inter-omics correlations) (Figure 2A, lines). Indeed, among the top features changing in each omics (Figure 2E), several were strongly correlated (|r| > 0.9) with cell type changes. For instance, 13 of the 28 cell types linked with rejuvenation showed a correlation between changes in cell composition and changes in the level of BPIFA1 (BPI Fold Containing Family A Member 1), a critical component of the innate immune response that prevents upper airway diseases. Similarly, changes in the levels of PC(38:3), AC(8:1) and HBG1 were correlated with composition changes in 13, 9, and 6 cell types, respectively. Overall, these results show that the modulation of cell type composition and proteomic changes associated with immunosenescence largely drive the biological age effects induced by TPE + IVIG intervention.

### Clinical markers are predictive of TPE + IVIG response

One important aspect of interventional studies is the ability to determine *a priori* which individuals will respond to treatment. In our study, this corresponds to individuals who displayed a decreased age acceleration after the treatments. We obtained omics data from individuals at baseline (before any treatment), and we wondered if the magnitude of the rejuvenation effects correlated with the levels of any clinical or omics markers before the treatment. We evaluated the correlation between the age-adjusted age acceleration difference at time point 2 and the levels of clinical and omics markers at baseline (Figure 3A). In addition to the omic markers, we evaluated 57 clinical markers, which are more accessible and practical to measure in a clinical setting. We found a significant correlation between 16 (28%) of these markers and the age acceleration differences (Figure 3B). High baseline levels of circulating bilirubin, glucose and alanine aminotransferase (ALT), which can signal liver dysfunction and metabolic disturbances^37–39^, were linked with the largest rejuvenation effects, suggesting that individuals with poorer health may experience the most significant improvements.

**Figure 3.**
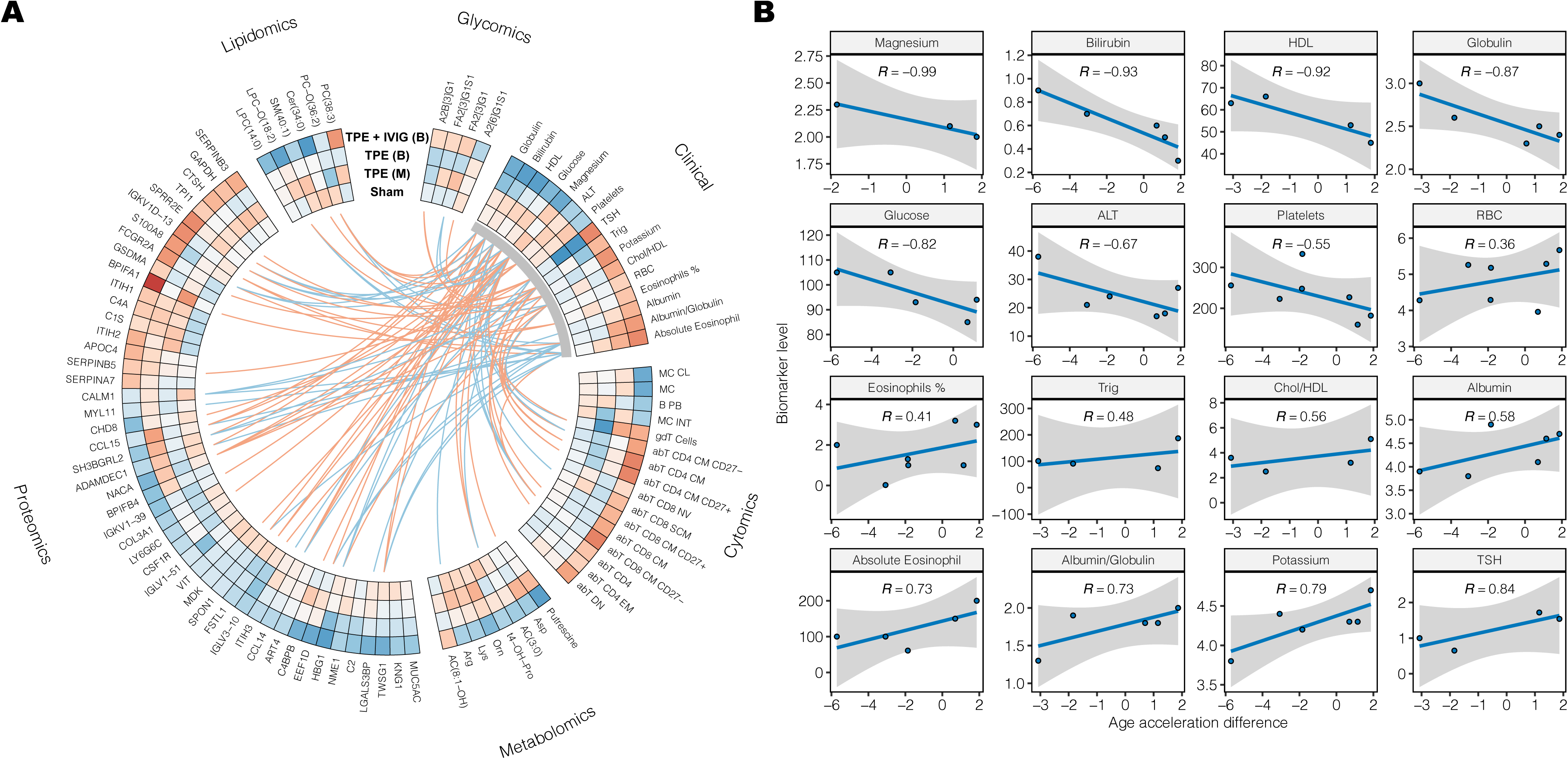
Baseline ‘omics’ and clinical biomarkers predict response to TPE-IVIG. A) ‘Omics’ features and clinical baseline levels correlated with the biological age response to TPE + IVIG. The lines connecting the features at the center indicate inter-feature correlations between clinical markers and other omics. B) Clinical markers significantly correlated with the rejuvenation effects of TPE + IVIG.

## Discussion

In this study, we aimed to assess the efficacy of TPE +/- IVIG in reducing BA and used multi-omics profiling to identify markers associated with the BA changes. Our findings demonstrate that three sessions of TPE treatment are sufficient to reduce biological age by 1.32 years, and that IVIG supplementation doubles the effect to 2.61 years. Previous studies have assessed biological age upon TPE treatment in humans and animal models using clinical markers^40^, pro-inflammatory markers^30^, and functional tests^41–43^. However, besides two ongoing clinical trials (ClinicalTrials.gov identifiers NCT05004220, NCT03353597), this is the first study in humans to report a decrease in biological age by TPE using well-validated epigenetic metrics of biological age.

We analyzed different types of epigenetic clocks and found that Systems age clocks^33^ displayed the most sensitive measures to the treatments, with “immune age” nearly decreasing 9.7 years after TPE + IVIG treatment. Consistently, we linked the rejuvenation effects to the restoration of immune cell composition changes characteristic of aging, as well as changes in the abundance of proteins linked to the immune response. This confirms our previously reported restoration of leukocyte subsets based on the expression of cell markers in PBMCs after TPE sessions and highlights the potential of TPE + IVIG therapy to significantly rejuvenate biological age by reversing immunosenescence.

We find that the subsequent therapy sessions restored most of the positive effects in BA induced by the first 3 sessions, with the restoration being linearly dependent on the initial BA effects. Previous studies have shown that TPE treatment could induce protein levels and cell composition to change beyond baseline levels^44,45^. We postulate that this may be due to either cell-intrinsic programming, which means that cells have inherent mechanisms to compensate and adapt to changes induced by the therapy, or exposome effects, in which lifestyle and environmental factors restore the body back to its baseline state, reducing the response to treatment over time. Additional analyses in larger cohorts using metabolomics as a proxy for the exposome are needed to address this hypothesis.

Interestingly, we discovered that proteins with correlated expression and rejuvenation effects tended to display increased abundance in senescent fibroblasts. While this may be unexpected for proteins showing increasing levels at a greater BA decrease, recent evidence from mice and other model organisms suggests that senescence not only may induce detrimental effects but is also important for tissue regeneration, wound healing, and tumor suppression^46^.

Our study also shows that baseline clinical testing can stratify patients for improved treatment efficacy. Here, whereas all subjects in this study were generally healthy and had laboratory values within the normal range, individuals with levels indicative of poorer health status, such as high blood glucose, elevated bilirubin and globulin and lower circulating albumin and potassium, benefit the most to TPE+IVIG treatment. However, the sample size of this study did not allow for the training and testing of predictive models to predict which subjects are likely to respond positively to the therapy. The failure to respond to TPE treatment has been observed in the treatment of different diseases^47,48^, and previous interventions have shown it to benefit unhealthy individuals the most^49^. This finding aligns with the rebound effects we observed in individuals with better health who may induce faster compensatory mechanisms.

Future research is necessary to further validate and optimize the rejuvenation effects of TPE, particularly when supplemented with IVIG. Long-term follow-up studies are necessary to determine the duration of the observed rejuvenation effects and the optimal treatment regime to minimize the rebound effects in BA we observed in this study. Additionally, larger sample sizes are required to enable the training and validation of predictive models that can identify the individuals most likely to benefit from TPE.

## Methods

### Participants and settings

Eligible patients were men and women over 50 years of age, except for one woman in her 40s, who had no known chronic clinical conditions. A subset of patients had clinical blood tests completed at Quest Labs (Bay Area, CA) or Life Extension Foundation Labs (online). Exclusion criteria included poor peripheral vascular access, diagnosis of active malignancy or active infection, late-stage Alzheimer’s disease, symptomatic coronary artery disease, congestive heart failure, restrictive pulmonary disease, asthma, taking growth hormones, stem cells, stem cell products or having a psychiatric disorder.

### Study Design

TPE was performed using a centrifugal blood separator (Spectra Optia, Lakewood, CO). During each procedure, one plasma volume was removed and replaced with 5% Albumin. The patients in the TPE-IVIG group received 2 gm of IVIG immediately after the TPE procedure. Enrolled patients were randomly allocated to four groups (in a 1:1:1:1 scheme): three TPE treatment and one control group that underwent a simulated TPE treatment through a noninvasive procedure (sham) that mimicked TPE but without any actual fluid replacement with the patient receiving approximately 250 cc of normal saline. The three TPE groups included a twice a week treatment once per month, a twice a week treatment once per month with IVIG added and a once a week once per month treatment. Samples were taken before the first, fourth and sixth TPE treatment. From a total of 240 TPE procedures, only 1 (0.42%) mild allergic reaction to albumin was observed. Samples were sent to each respective core facility or third-party company for processing and analysis. Patients, caregivers, and raters were blinded.

Subjects in the Sham group had a peripheral vein IV inserted in each arm, similar to the actual treatment subjects, but with only normal saline IV fluid. Dark curtains were used to hide the apheresis device and the pumps from the participants’ field of view. The apheresis device was turned on and ran using a container filled with water, with both the access and the return lines submerged in the container to simulate blood flow through the machine. The device was programmed using the patient’s information (height, weight and etc) and rinseback procedure was done to maintain the perception that the actual TPE procedure was finished.

### Epigenomics

DNA methylation was evaluated using TruAge (developed by TruDiagnostic Inc., Lexington, KY). Peripheral whole blood samples were obtained and then mixed with a lysis buffer to preserve the cells. DNA extraction was performed, and 500 ng of DNA were subjected to bisulfite conversion using the EZ DNA Methylation Kit from Zymo Research, following the manufacturer’s protocol. The bisulfite-converted DNA samples were randomly allocated to designated Illumina Infinium EPIC850k Beadchip wells. The samples were amplified, hybridized onto the array, and subsequently stained. After washing steps, the variety was imaged using the Illumina iScan SQ instrument to capture raw image intensities, enabling further analysis. Raw IDAT files were processed using the minfi pipeline^50^. Low-quality samples were detected using ENMix by examining the variance of internal controls and flagging those with values more than 3 standard deviations from the mean control probe value^51^. However, no outlier samples were found, so all samples were included in the analysis. Single-sample Noob (ssNoob) normalization was used in order to consistently normalize the samples across the multiple array types. The algorithms analyzed by TruAge include first-(Horvath and Hannum) and second-(phenoAge, systemsAge, OMICAge, and GrimAge). Epigenetic age acceleration (EAA) of the first and second generation clocks was calculated as the residual of each clock regressed upon chronological age. To account for any perceived batch effects, the first 3 principal components calculated from the technical probes were calculated and used as adjustment factors when calculating the EAA.

### Proteomics

All water, methanol, acetonitrile, formic acid, and trifluoracetic acid used were LC/MS grade. Perchloric acid (70%) was purchased from Sigma Aldrich (311421-50ML). Sequencing grade modified trypsin was obtained from Promega (V511B). Indexed retention time standards (iRT peptides) were purchased from Biognosys. HLB columns (1cc 10 mg) were purchased from Waters (186000383).

Plasma samples were thawed on ice, and two 125 µL aliquots per sample were transferred to 1.5 mL tubes. Samples were diluted 10-fold with water and mixed with 70% perchloric acid to a final concentration of 3.5%^52–54^. Samples were incubated at -20 °C for 15 min and subsequently centrifuged at 3,200 xg for 60 min at 4 °C. The supernatant was transferred to new tubes and acidified with 1% trifluoracetic acid. Perchloric acid was removed using HLB columns (10 mg, Waters). Columns were conditioned with 800 µL methanol and washed with 1,600 µL of 0.1% trifluoracetic acid (TFA). The acidified supernatant was loaded onto the column, washed with 2,400 µL of 0.1% TFA, and proteins were eluted with 800 µL of 0.1% TFA in 90% acetonitrile. Eluates were dried using a speed-vac and resuspended in 50 µL of 0.5% sodium dodecyl sulfate in 100 mM triethylammonium bicarbonate (TEAB).

Proteins were denatured with 10% SDS at 90°C for 10 min, cooled to room temperature, and pH adjusted to ∼7 using 1M TEAB. Proteins were reduced with 250 mM dithiothreitol at 56 °C for 10 min, alkylated with 250 mM iodoacetamide in the dark for 30 min, and acidified with 12% phosphoric acid. Proteins were trapped and digested on S-Trap mini spin columns according to the manufacturer’s instructions. Briefly, reduced and alkylated proteins were diluted with S-Trap buffer, loaded onto the column, and digested with 4 µg trypsin in 50 mM TEAB in two stages - 1 h at 47 °C followed by additional 4 µg trypsin and overnight incubation at 37 °C. Peptides were eluted sequentially with 50 mM TEAB, 0.5% formic acid, and 0.5% formic acid in 50% acetonitrile. Eluates were pooled, dried using a speed-vac, and resuspended in 1% formic acid. Peptides were desalted using HLB columns (10 mg, Waters). Columns were conditioned with 0.2% formic acid in 50% acetonitrile and washed with 0.2% formic acid. Samples were loaded, washed with 0.2% formic acid, and eluted with 0.2% formic acid in 50% acetonitrile. Eluates were dried using a speed-vac and resuspended in 100 µL 0.2% formic acid. Samples were stored at -20 °C until analysis. Prior to LC-MS/MS analysis, peptides were diluted 1:1 with 0.2% formic acid and spiked with 0.5 µL of iRT peptides (Biognosys)^55^.

Reverse-phase HPLC-MS/MS data was acquired using a Waters M-Class HPLC (Waters, Massachusetts, MA) connected to a ZenoTOF 7600 (SCIEX, Redwood City, CA) with an OptiFlow Turbo V Ion Source (SCIEX) equipped with a microelectrode^56^. The chromatographic solvent system consisted of 0.1% formic acid in water (solvent A) and 99.9% acetonitrile, 0.1% formic acid in water (solvent B). Digested peptides (4 µL) were loaded onto a Luna Micro C18 trap column (20 x 0.30 mm, 5 µm particle size; Phenomenex, Torrance, CA) over a period of 2 minutes at a flow rate of 10 µL/min using 100% solvent A. Peptides were eluted onto a Kinetex XB-C18 analytical column (150 x 0.30 mm, 2.6 µm particle size; Phenomenex) at a flow rate of 5 µL/min using a 120 minute microflow gradient (as described below), with each gradient ranging from 5 to 32% solvent B. Briefly, 4 µL of digested peptides were loaded at 5% B and separated using a 120 minute linear gradient from 5 to 32% B, followed by an increase to 80% B for 1 minute, a hold at 80% B for 2 minutes, a decrease to 5% B for 1 minute, and a hold at 5% B for 6 minutes. The total HPLC acquisition time was 130 minutes. The following MS parameters were used for all acquisitions: ion source gas 1 at 10 psi, ion source gas 2 at 25 psi, curtain gas at 30 psi, CAD gas at 7 psi, source temperature at 200°C, column temperature at 30°C, polarity set to positive, and spray voltage at 5000V.

All human samples were acquired in data-independent acquisition mode (DIA) analysis with two technical replicates for each biological sample replicate^57–59^. Briefly, the DIA-MS method on the ZenoTOF 7600 system is comprised of A survey MS1 scan (mass range: 395-1005 m/z), with an accumulation time of 100 ms, a declustering potential of 80 V, and a collision energy of 10 V. MS2 scans were acquired using 80 variable width windows across the precursor ion mass range (399.5-1000.5 m/z), with an MS2 accumulation time of 25 ms, dynamic collision energy enabled, charge state 2 selected, and Zeno pulsing enabled (total cycle time 2.5 seconds).

All data files were processed with Spectronaut v16 (version 16.0.220524.5300; Biognosys) performing a directDIA search using the UniProt *Homo sapiens* reference proteome with 20,423 entries, accessed on 06/30/2023. Dynamic data extraction parameters and precision iRT calibration with local non-linear regression were used. Trypsin/P was specified as the digestion enzyme, allowing for specific cleavages and up to two missed cleavages. Methionine oxidation and protein N-terminus acetylation were set as dynamic modifications, while carbamidomethylation of cysteine was set as a static modification. Protein group identification (grouping for protein isomers) required at least 2 unique peptides and was performed using a 1% q-value cutoff for both the precursor ion and protein level. Protein quantification was based on the peak areas of extracted ion chromatograms (XICs) of 3 – 6 MS2 fragment ions, specifically b- and y-ions, with automatic normalization and 1% q-value data filtering applied. Relative protein abundance changes were compared using the Storey method with paired t-tests and p-values corrected for multiple testing using group wise testing corrections^60^.

### Glycomics

Glycan data was derived as described previously^61^. For the analytical precision analysis, AMC and long-term variability, the IgG isolation, IgG *N*-glycan release and labeling were performed using a Genos-Glycanage IgG glycome profiling kit (Genos, Osijek, Croatia) and subsequent capillary gel electrophoresis with laser-induced fluorescence (CGE-LIF) analysis was adapted from previously published protocols^62,63^. The process of extracting IgG involved diluting 25 µL of subject plasma samples and three distinct plasma standards in quadruplicate, serving as technical replicates of a known, previously analyzed, glycome. This dilution was carried out using a 1:7 ratio with a 1 × PBS buffer, which was prepared in-house. Additionally, blank samples containing ultrapure water, without any analyte, were included to monitor and control for potential cross-contamination. The diluted samples were resuspended and filtered through a wwPTFE filter plate with 0.45-µm pore size (Pall corporation, New York, NY, USA) using a vacuum manifold and pump (Pall corporation, New York, NY, USA). The filtered samples were transferred to a CIM® r-Protein G LLD 0.05 mL monolithic 96-well plate (Sartorius BIA Separations, Ajdovščina, Slovenia), where they underwent binding and subsequent washing steps with phosphate-buffered saline (1 × PBS) buffer (0.25 M NaCl, increased ionic strength, prepared in-house). Elution of the bound IgG was achieved by employing 0.1 M formic acid neutralized with ammonium bicarbonate buffer (Sigma-Aldrich, St. Louis, MO, USA). The eluted IgG fraction (20 µL) was dried and prepared for the subsequent steps in the protocol.

The dried IgG samples were consecutively treated with 1.66 × PBS, 0.5% sodium dodecyl sulfate (SDS) and 2% Igepal (Sigma-Aldrich, St. Louis, MO, USA/Invitrogen Thermo Fisher Scientific, Carlsbad, CA, USA), to denature the IgG, followed by incubation with 1.2U of the enzyme PNGase F (Promega, Madison, WI, USA) at 37°C for 3 h to release its *N*-glycans. The released glycans were then labeled by mixing APTS (8-aminopyrene-1,3,6-trisulfonic acid) (Synchem, Felsberg, Germany) fluorescent dye with the reducing agent 2-picoline borane (Sigma-Aldrich, St. Louis, MO, USA) and subjected to a 16-h incubation at 37 °C.

After incubation, the labeling reaction was halted by the addition of 80% acetonitrile (ACN, Carlo Erba, Milan, Italy). The clean-up of the released fluorescently labeled IgG *N*-glycans was conducted using solid-phase extraction utilizing Bio-Gel P-10 as a hydrophilic stationary phase. The entire sample volume was transferred to the filter plate containing the Bio-Gel P-10. The excess label and reducing agent were removed by five washes with 80% ACN/100 mM triethylamine (Sigma-Aldrich, St. Louis, MO, USA), followed by three washes with 80% ACN. Finally, APTS labeled IgG *N*-glycans were eluted in ultra-pure water.

For CGE-LIF analysis, 3 µL of purified IgG *N*-glycans combined with 7 µL of Hi-Di Formamide were analyzed using an ABI3500 Genetic Analyzer (Thermo Fisher Scientific, Waltham, MA, USA) equipped with a 50-cm long 8-capillary array filled with POP-7 polymer as a separation matrix. Run parameters were set as follows: run time 1000 s, injection time 12 s, injection voltage 15 kV, run voltage 15 kV, and oven temperature 60 °C. The resulting electropherograms were manually integrated into 27 glycan peaks using the Empower 3 software (Waters, Milford, MA, USA). The amount of glycan structures in a peak was expressed as a percentage of the total integrated area (total area normalization). In addition, six derived glycan traits were calculated for glycans with shared structural features^64,65^.

### Metabolomics and Lipidomics

Targeted metabolomics was performed on plasma samples utilizing the Biocrates AbsoluteIDQ p400 HR kit. Prior to analysis, a system suitability test and instrument calibration were performed with the QExactive mass spectrometer (Thermo Scientific). The experimental procedure involved processing plasma samples, blanks, calibration standards (7-point), and quality controls according to the manufacturer’s recommendations.

Specifically, 10 µL of plasma was added to a pre-loaded filter plate containing internal standards and dried using ultra-pure nitrogen. Subsequent derivatization with 5% phenylisothiocyanate in pyridine, ethanol, and water (in a 1:1:1 ratio), followed by extraction with 5 mM ammonium acetate in methanol, yielded extracts that were collected into a 96-deep well plate via centrifugation. Mass spectrometric analysis was conducted using the Thermo QExactive mass spectrometer in positive ionization mode. Chromatographic separation utilized a proprietary Biocrates column, employing 0.2% formic acid in water as buffer A and 0.2% formic acid in acetonitrile as buffer B.

For the analysis of lipids, acylcarnitines, and hexoses, flow injection analysis (FIA) was employed without a column, utilizing the Biocrates FIA additive mobile phase. Data processing and lipid quantification were performed using Thermo Excalibur, QuanBrowser, and MetIDQ software. Normalization of peak areas corresponding to metabolites was conducted relative to their respective internal standards. Target metabolite concentrations were estimated linearly based on observed concentrations in quality control samples, and a seven-point quadratic calibration approach was implemented where applicable. All chemicals and solvents used were LCMS grade.

### Cytomics

Blood was collected into lavender-top vacutainer tubes (K-EDTA, BD) and kept on ice until processing the same day. Peripheral blood mononuclear cells (PBMC) were prepared from the whole blood by density gradient centrifugation using Ficoll-Paque PLUS (Cytivia) according to the package directions, followed by lysis of residual erythrocytes with ACK buffer (Gibco). After washing twice in dPBS, PBMC were cryopreserved in 90% fetal bovine serum (FBS) and 10% DMSO and stored in liquid nitrogen until analysis.

Vials of frozen PBMC (∼2 million) were thawed rapidly by swirling in a 37°C water bath and immediately transferred to 15mL conical tubes with 10 mL of pre-warmed RPMI medium with 10% FBS, then pelleted for 5 minutes at 300g in a benchtop TC centrifuge. After removal of the supernatant, the cells were resuspended in 4 mL of warmed RPMI/10% FBS and allowed to recover in a TC incubator for three hours prior to staining. After recovery, the cells were divided equally for analysis with the SPiDERGal and intracellular reagent panels. For the SPiDERGal workflow, the cells were left in medium and treated with 1 μM Bafilomycin A1 (Abcam) for 1 hour, followed by 667 nM of SPiDERGal (ThermoFisher) for one additional hour, while the cells for the IC panel were pelleted and kept on ice for staining. For both panels, the cells were resuspended in 200 µL of dPBS in V-bottom plates and incubated for 20 minutes on ice with 1X Live/Dead Blue (ThermoFisher) plus 5 μg/mL of human IgG (Sigma). Surface marker-specific antibodies plus FBS to 2% v/v were then added (as specified in the attached table), and the cells were incubated on ice in the dark for 1 hour, then washed once in PBS/2% FBS. For the SPiDER panel, the cells were then taken up in 150µL of PBS/2% FBS and acquired. For the IC panel, the cells were taken up in 150μL of FOXP3 Fix/perm buffer (ThermoFisher) and incubated on ice for 20 minutes, pelleted, washed once with FOXP3 Perm-wash buffer (ThermoFisher), and then stained with the IC marker-specific antibodies in Perm-wash buffer as specified in the attached table. They were then pelleted, washed once in Perm-wash buffer, and resuspended in PBS/2% FBS for acquisition.

The stained PBMC were acquired on a 5-laser Cytek Aurora spectral flow cytometer (Cytek), and the raw data spectrally unmixed using the SpectroFlo software package (Cytek). Subsequent correction of spectral compensation and manual gating to identify populations were performed using FlowJo (software (BD), and population frequencies (as % of Parent pop and percent of live leukocytes) and numbers were tabulated for subsequent bioinformatic analysis.

### Bioinformatic Analysis

Using the Epigenetic age acceleration (EAA) calculated (see Methods - Epigenomics) we calculated the age acceleration difference as:

Age acceleration difference at time point 2 = EAA (time point 2) - EAA (time point 1)

Age acceleration difference at time point 3 = EAA (time point 3) - EAA (time point 1)

The mean age acceleration difference across individuals was calculated for each of the 35 different epigenetic clocks in each group. The average age acceleration difference for the epigenetic clocks was compared between treatment groups and sham using the Wilcoxon test.

Changes in feature levels in each omics were calculated for time points 2 or 3 versus 1 and correlated against the age acceleration difference at time points 2 and 3. For down-stream analysis, we only considered features measured in at least 3 samples. Correlations were transformed into z-scores using Fisher transformation and compared against zero and sham using a t-test. P-values were adjusted for multiple tests in each omics using Benjamini-Hochberg method^66^. Features with FDR < 0.05 against zero and sham were considered statistically significant. In the case of the comparison against baseline levels of clinical and omics markers, the correlations were calculated between the baseline levels and age acceleration difference and the same method was used to calculate the statistical significance. Circular and rectangular heatmaps were generated using the package circlize and ComplexHeatmap^47^.

Gene ontology enrichment analysis was performed using the function enrichGO implemented in the package clusterProfiler^67^. Only GO biological process terms with a set size between 50 and 500 were analyzed. Enrichment results were plotted using the package CellPlot^68^. Data from the SASP Atlas was obtained from www.saspatlas.com^36^. Proteins with q-value < 0.05 were considered in the comparative analysis against markers of rejuvenation.

## Data Availability

The raw data and complete mass spectrometry data sets have been uploaded to the Mass Spectrometry Interactive Virtual Environment (MassIVE) repository, which is maintained by the Center for Computational Mass Spectrometry at the University of California San Diego. Raw data and complete MS data sets have been uploaded to the MassIVE repository and can be downloaded using the following link: https://massive.ucsd.edu/ProteoSAFe/dataset.jsp?task=945e8acb15054ae895937c3b84d926b9 (MassIVE ID number: MSV000095275; ProteomeXchange ID: PXD053805).

## Acknowledgments

This study was funded by Circulate, Inc.

## Bibliography

1. Ageing and health. Accessed July 1, 2024. https://www.who.int/news-room/fact-sheets/detail/ageing-and-health

2. Sigmond T, Vellai T. Lysosomal alteration links food limitation to longevity. Nat Aging. 2023;3(9):1048–1050. doi:10.1038/s43587-023-00483-1

3. Sayed N, Huang Y, Nguyen K, et al. An inflammatory aging clock (iAge) based on deep learning tracks multimorbidity, immunosenescence, frailty and cardiovascular aging. Nat Aging. 2021;1(7):598–615. doi:10.1038/s43587-021-00082-y

4. Meier TB, Huber DL, Goeckner BD, et al. Association of Blood Biomarkers of Inflammation With Acute Concussion in Collegiate Athletes and Military Service Academy Cadets. Neurology. 2024;102(2):e207991. doi:10.1212/WNL.0000000000207991

5. Waziry R, Gras L, Sedaghat S, et al. Quantification of biological age as a determinant of age-related diseases in the Rotterdam Study: a structural equation modeling approach. Eur J Epidemiol. 2019;34(8):793–799. doi:10.1007/s10654-019-00497-3

6. Goeminne LJE, Eames A, Tyshkovskiy A, et al. Plasma-based organ-specific aging and mortality models unveil diseases as accelerated aging of organismal systems. Published online April 10, 2024:2024.04.08.24305469. doi:10.1101/2024.04.08.24305469

7. Belsky DW, Caspi A, Corcoran DL, et al. DunedinPACE, a DNA methylation biomarker of the pace of aging. Deelen J, Tyler JK, Suderman M, Deelen J, eds. eLife. 2022;11:e73420. doi:10.7554/eLife.73420

8. Furman D, Campisi J, Verdin E, et al. Chronic inflammation in the etiology of disease across the life span. Nat Med. 2019;25(12):1822–1832. doi:10.1038/s41591-019-0675-0

9. Bell CG, Lowe R, Adams PD, et al. DNA methylation aging clocks: challenges and recommendations. Genome Biol. 2019;20(1):249. doi:10.1186/s13059-019-1824-y

10. Johnson AA, Shokhirev MN, Wyss-Coray T, Lehallier B. Systematic review and analysis of human proteomics aging studies unveils a novel proteomic aging clock and identifies key processes that change with age. Ageing Res Rev. 2020;60:101070. doi:10.1016/j.arr.2020.101070

11. Hertel J, Friedrich N, Wittfeld K, et al. Measuring Biological Age via Metabonomics: The Metabolic Age Score. J Proteome Res. 2016;15(2):400–410. doi:10.1021/acs.jproteome.5b00561

12. Rutledge J, Oh H, Wyss-Coray T. Measuring biological age using omics data. Nat Rev Genet. 2022;23(12):715–727. doi:10.1038/s41576-022-00511-7

13. Alpert A, Pickman Y, Leipold M, et al. A clinically meaningful metric of immune age derived from high-dimensional longitudinal monitoring. Nat Med. 2019;25(3):487–495. doi:10.1038/s41591-019-0381-y

14. Yaskolka Meir A, Keller M, Bernhart SH, et al. Lifestyle weight-loss intervention may attenuate methylation aging: the CENTRAL MRI randomized controlled trial. Clin Epigenetics. 2021;13(1):48. doi:10.1186/s13148-021-01038-0

15. Fitzgerald KN, Campbell T, Makarem S, Hodges R. Potential reversal of biological age in women following an 8-week methylation-supportive diet and lifestyle program: a case series. Aging. 2023;15(6):1833–1839. doi:10.18632/aging.204602

16. Fitzgerald KN, Hodges R, Hanes D, et al. Potential reversal of epigenetic age using a diet and lifestyle intervention: a pilot randomized clinical trial. Aging. 2021;13(7):9419–9432. doi:10.18632/aging.202913

17. Fiorito G, Caini S, Palli D, et al. DNA methylation-based biomarkers of aging were slowed down in a two-year diet and physical activity intervention trial: the DAMA study. Aging Cell. 2021;20(10):e13439. doi:10.1111/acel.13439

18. Chen L, Dong Y, Bhagatwala J, Raed A, Huang Y, Zhu H. Effects of Vitamin D3 Supplementation on Epigenetic Aging in Overweight and Obese African Americans With Suboptimal Vitamin D Status: A Randomized Clinical Trial. J Gerontol Ser A. 2019;74(1):91–98. doi:10.1093/gerona/gly223

19. Vetter VM, Spira D, Banszerus VL, Demuth I. Epigenetic Clock and Leukocyte Telomere Length Are Associated with Vitamin D Status but not with Functional Assessments and Frailty in the Berlin Aging Study II. J Gerontol Ser A. 2020;75(11):2056–2063. doi:10.1093/gerona/glaa101

20. Vetter VM, Sommerer Y, Kalies CH, Spira D, Bertram L, Demuth I. Vitamin D supplementation is associated with slower epigenetic aging. GeroScience. 2022;44(3):1847–1859. doi:10.1007/s11357-022-00581-9

21. Pavanello S, Campisi M, Tona F, Dal Lin C, Iliceto S. Exploring Epigenetic Age in Response to Intensive Relaxing Training: A Pilot Study to Slow Down Biological Age. Int J Environ Res Public Health. 2019;16(17):3074. doi:10.3390/ijerph16173074

22. Li M, Bao L, Zhu P, Wang S. Effect of metformin on the epigenetic age of peripheral blood in patients with diabetes mellitus. Front Genet. 2022;13. doi:10.3389/fgene.2022.955835

23. Moqri M, Herzog C, Poganik JR, et al. Biomarkers of Aging for the Identification and Evaluation of Longevity Interventions. Cell. 2023;186(18):3758–3775. doi:10.1016/j.cell.2023.08.003

24. Clement J, Yan Q, Agrawal M, et al. Umbilical cord plasma concentrate has beneficial effects on DNA methylation GrimAge and human clinical biomarkers. Aging Cell. 2022;21(10):e13696. doi:10.1111/acel.13696

25. Abel JJ, Rowntree LG, Turner BB. Plasma Removal with Return of Corpuscles (plasmaphaeresis): First Paper. J Pharmacol Exp Ther. 1914;5(6):625–641.

26. Solomon A, Fahey JL. Plasmapheresis therapy in macroglobulinemia. Ann Intern Med. 1963;58:789–800. doi:10.7326/0003-4819-58-5-789

27. Spectra Optia Apheresis System with the Depuro D2000 - Letter of Authorization | FDA. Accessed June 23, 2024. https://www.fda.gov/media/136834

28. Kiprov DD. A Paradigm Shift in the Utilization of Therapeutic Plasmapheresis in Clinical Practice. 12(5).

29. Radhakrishnan K, Thokala RP, Anandan A, Rengan CS. Role of therapeutic plasma exchange in rat killer (yellow phosphorous) poisoning. Asian J Transfus Sci. 2023;17(1):74. doi:10.4103/ajts.ajts_20_21

30. Kim D, Kiprov DD, Luellen C, et al. Old plasma dilution reduces human biological age: a clinical study. GeroScience. 2022;44(6):2701–2720. doi:10.1007/s11357-022-00645-w

31. Megha KB, Mohanan PV. Role of immunoglobulin and antibodies in disease management. Int J Biol Macromol. 2021;169:28–38. doi:10.1016/j.ijbiomac.2020.12.073

32. Katragkou A, Roilides E, Walsh TJ. Role of Immunoglobulin Therapy to Prevent and Treat Infections. In: Segal BH, ed. Management of Infections in the Immunocompromised Host. Springer International Publishing; 2018:339–358. doi:10.1007/978-3-319-77674-3_17

33. Sehgal R, Markov Y, Qin C, et al. Systems Age: A single blood methylation test to quantify aging heterogeneity across 11 physiological systems. Published online May 28, 2024:2023.07.13.548904. doi:10.1101/2023.07.13.548904

34. Egorov ES, Kasatskaya SA, Zubov VN, et al. The Changing Landscape of Naive T Cell Receptor Repertoire With Human Aging. Front Immunol. 2018;9. doi:10.3389/fimmu.2018.01618

35. Ligotti ME, Aiello A, Accardi G, et al. Analysis of T and NK cell subsets in the Sicilian population from young to supercentenarian: The role of age and gender. Clin Exp Immunol. 2021;205(2):198–212. doi:10.1111/cei.13606

36. Basisty N, Kale A, Jeon OH, et al. A proteomic atlas of senescence-associated secretomes for aging biomarker development. PLOS Biol. 2020;18(1):e3000599. doi:10.1371/journal.pbio.3000599

37. Hamoud AR, Weaver L, Stec DE, Hinds TD. Bilirubin in the Liver-Gut Signaling Axis. Trends Endocrinol Metab TEM. 2018;29(3):140–150. doi:10.1016/j.tem.2018.01.002

38. Güemes M, Rahman SA, Hussain K. What is a normal blood glucose? Arch Dis Child. 2016;101(6):569–574. doi:10.1136/archdischild-2015-308336

39. Liu Z, Que S, Xu J, Peng T. Alanine aminotransferase-old biomarker and new concept: a review. Int J Med Sci. 2014;11(9):925–935. doi:10.7150/ijms.8951

40. Li X, Zhang J, Sun C, et al. Application of biological age assessment of Chinese population in potential anti-ageing technology. Immun Ageing A. 2018;15:33. doi:10.1186/s12979-018-0140-9

41. Mehdipour M, Amiri P, Liu C, et al. Small-animal blood exchange is an emerging approach for systemic aging research. Nat Protoc. 2022;17(11):2469–2493. doi:10.1038/s41596-022-00731-5

42. Mehdipour M, Mehdipour T, Skinner CM, et al. Plasma dilution improves cognition and attenuates neuroinflammation in old mice. GeroScience. 2021;43(1):1–18. doi:10.1007/s11357-020-00297-8

43. Mehdipour M, Skinner C, Wong N, et al. Rejuvenation of three germ layers tissues by exchanging old blood plasma with saline-albumin. Aging. 2020;12(10):8790–8819. doi:10.18632/aging.103418

44. Foettinger F, Pilz G, Wipfler P, et al. Immunomodulatory Aspects of Therapeutic Plasma Exchange in Neurological Disorders—A Pilot Study. Int J Mol Sci. 2023;24(7):6552. doi:10.3390/ijms24076552

45. Orme JJ, Enninga EAL, Lucien-Matteoni F, et al. Therapeutic plasma exchange clears circulating soluble PD-L1 and PD-L1-positive extracellular vesicles. J Immunother Cancer. 2020;8(2):e001113. doi:10.1136/jitc-2020-001113

46. de Magalhães JP. Cellular senescence in normal physiology. Science. 2024;384(6702):1300–1301. doi:10.1126/science.adj7050

47. Aradillas E, Schwartzman RJ, Grothusen JR, Goebel A, Alexander GM. Plasma Exchange Therapy in Patients with Complex Regional Pain Syndrome. Pain Physician. 2015;18(4):383–394.

48. Franke D, Zimmering M, Wolfish N, Ehrich JHH, Filler G. Treatment of FSGS with plasma exchange and immunadsorption. Pediatr Nephrol. 2000;14(10):965–969. doi:10.1007/PL00013420

49. Dioum EHM, Schneider KL, Vigerust DJ, et al. Oats Lower Age-Related Systemic Chronic Inflammation (iAge) in Adults at Risk for Cardiovascular Disease. Nutrients. 2022;14(21):4471. doi:10.3390/nu14214471

50. Aryee MJ, Jaffe AE, Corrada-Bravo H, et al. Minfi: a flexible and comprehensive Bioconductor package for the analysis of Infinium DNA methylation microarrays. Bioinformatics. 2014;30(10):1363–1369. doi:10.1093/bioinformatics/btu049

51. Xu Z, Niu L, Taylor JA. The ENmix DNA methylation analysis pipeline for Illumina BeadChip and comparisons with seven other preprocessing pipelines. Clin Epigenetics. 2021;13(1):216. doi:10.1186/s13148-021-01207-1

52. Winzler RJ, Devor AW. Studies on the mucoproteins of human plasma; determination and isolation. J Clin Invest. 1948;27(5):609–616.

53. Zougman A, Wiśniewski JR. Beyond Linker Histones and High Mobility Group Proteins: Global Profiling of Perchloric Acid Soluble Proteins. J Proteome Res. 2006;5(4):925–934. doi:10.1021/pr050415p

54. Viode A, van Zalm P, Smolen KK, et al. A simple, time- and cost-effective, high-throughput depletion strategy for deep plasma proteomics. Sci Adv. 2023;9(13):eadf9717. doi:10.1126/sciadv.adf9717

55. Escher C, Reiter L, MacLean B, et al. Using iRT, a normalized retention time for more targeted measurement of peptides. Proteomics. 2012;12(8):1111–1121. doi:10.1002/pmic.201100463

56. Burton JB, Silva-Barbosa A, Bons J, et al. Substantial downregulation of mitochondrial and peroxisomal proteins during acute kidney injury revealed by data-independent acquisition proteomics. Proteomics. 2024;24(5):e2300162. doi:10.1002/pmic.202300162

57. Gillet LC, Navarro P, Tate S, et al. Targeted Data Extraction of the MS/MS Spectra Generated by Data-independent Acquisition: A New Concept for Consistent and Accurate Proteome Analysis. Mol Cell Proteomics MCP. 2012;11(6):O111.016717. doi:10.1074/mcp.O111.016717

58. Collins BC, Hunter CL, Liu Y, et al. Multi-laboratory assessment of reproducibility, qualitative and quantitative performance of SWATH-mass spectrometry. Nat Commun. 2017;8:291. doi:10.1038/s41467-017-00249-5

59. Schilling B, Gibson BW, Hunter CL. Generation of High-Quality SWATH® Acquisition Data for Label-free Quantitative Proteomics Studies Using TripleTOF® Mass Spectrometers. Methods Mol Biol Clifton NJ. 2017;1550:223–233. doi:10.1007/978-1-4939-6747-6_16

60. Burger T. Gentle Introduction to the Statistical Foundations of False Discovery Rate in Quantitative Proteomics. J Proteome Res. 2018;17(1):12–22. doi:10.1021/acs.jproteome.7b00170

61. Rapčan B, Song M, Frkatović-Hodžić A, et al. Glycan clock of ageing—analytical precision and time-dependent inter- and i-individual variability. GeroScience. Published online June 14, 2024. doi:10.1007/s11357-024-01239-4

62. Pučić M, Knežević A, Vidič J, et al. High Throughput Isolation and Glycosylation Analysis of IgG– Variability and Heritability of the IgG Glycome in Three Isolated Human Populations. Mol Cell Proteomics MCP. 2011;10(10):M111.010090. doi:10.1074/mcp.M111.010090

63. Hanić M, Lauc G, Trbojević-Akmačić I. N-Glycan Analysis by Ultra-Performance Liquid Chromatography and Capillary Gel Electrophoresis with Fluorescent Labeling. Curr Protoc Protein Sci. 2019;97(1):e95. doi:10.1002/cpps.95

64. Trbojević Akmačić I, Ugrina I, Štambuk J, et al. High-throughput glycomics: Optimization of sample preparation. Biochem Mosc. 2015;80(7):934–942. doi:10.1134/S0006297915070123

65. Gornik O, Wagner J, Pučić M, Knežević A, Redžić I, Lauc G. Stability of N-glycan profiles in human plasma. Glycobiology. 2009;19(12):1547–1553. doi:10.1093/glycob/cwp134

66. Benjamini Y, Hochberg Y. Controlling the False Discovery Rate: A Practical and Powerful Approach to Multiple Testing. J R Stat Soc Ser B Methodol. 1995;57(1):289–300. doi:10.1111/j.2517-6161.1995.tb02031.x

67. Wu T, Hu E, Xu S, et al. clusterProfiler 4.0: A universal enrichment tool for interpreting omics data. The Innovation. 2021;2(3):100141. doi:10.1016/j.xinn.2021.100141

68. dieterich-lab/CellPlot. Published online March 27, 2024. Accessed July 5, 2024. https://github.com/dieterich-lab/CellPlot

